# Mental health as a mediator of the association between educational inequality and cardiovascular disease: A Mendelian randomisation study

**DOI:** 10.1101/2020.09.10.20191825

**Authors:** Daniel P Jones, Robyn E Wootton, Dipender Gill, Alice R Carter, David Gunnell, Marcus R Munafò, Hannah M Sallis

## Abstract

**Background:** Education is inversely associated with cardiovascular disease. Several mediators for this association have been established but a significant proportion of the protective effect remains unaccounted for. Mental health is a proposed mediator, but current evidence is mixed and subject to bias from confounding factors and reverse causation. Mendelian randomisation (MR) is an instrumental variable technique that uses genetic proxies for exposures and mediators to reduce such bias.

**Methods and Results:** We used logistic regression and two-step MR analyses to investigate whether educational attainment affects risk of mental health disorders. We then performed observational and MR mediation analyses to explore whether mental health disorders mediate the association between educational attainment and risk of cardiovascular disease. Higher levels of educational attainment were associated with reduced depression, anxiety and cardiovascular disease in observational analyses [Odds Ratio (95% Confidence interval) 0.79 (0.77-0.81), 0.76 (0.73-0.79) and 0.79 (0.78-0.81) respectively], and MR analyses provided support for these reflecting causal effects [OR (95% CI) 0.72 (0.67-0.77), 0.50 (0.42-0.59) and 0.62 (0.58-0.66) respectively]. Both anxiety and depression were associated with cardiovascular disease in observational analyses [OR (95% CI) 1.63 (1.49-1.79) and OR (95% CI) 1.70 (1.59-1.82) respectively] but only depression was associated in the MR analyses [OR (95% CI) 1.09 (1.03-1.15)]. Roughly 6% of the total protective effect of education on cardiovascular disease was mediated by depression.

**Conclusions:** Higher levels of educational attainment protect against mental health disorders and reduced depression accounts for a small proportion of the total protective effect of education on cardiovascular disease.

## Introduction

Cardiovascular disease is a leading cause of global morbidity and mortality^1^. The association between socio-economic inequality and cardiovascular disease is well-established, with those living in deprived areas typically having much higher cardiovascular mortality than those in the less deprived areas^2^. More specifically, disparities in educational attainment have recently been shown to affect cardiovascular disease using both conventional and genetic epidemiological techniques^3^ In the latter, cardiovascular risk was estimated to decrease by a third for every 3.6 years of additional full-time education past the age of eleven ^4^.

A number of well-recognised cardiovascular risk factors appear to act as mediators for education^5^. For example, low educational attainment is associated with increased tobacco smoking, higher BMI, and higher blood pressure which in turn are linked to increased risk of cardiovascular disease^6^. However, even taking these mediators into account, there is a significant proportion of the association between education and cardiovascular disease that remains unaccounted for.

Existing evidence suggests an association between mental health and cardiovascular disease as well as an association between education and mental health^7,8^. Consequently, mental health may be another potential mediator between education and cardiovascular disease. For example, educational inequality may lead to disparities in psychological development by affecting exposure to stressors, resources to cope with stress, and negative external societal perception^9^. Poor mental health could, in turn, lead to increased cardiovascular disease risk with proposed mechanisms including chronic inflammation^10^, increased sympathetic nervous system activation^11^, and increased cortisol exposure^12^.

Conventional observational studies help identify and quantify any associations between the two that may be due to causal effects. Those that have explored education, mental health and cardiovascular disease have, so far, proved inconclusive^13,14^. Furthermore, observational evidence can be subject to bias from confounding and reverse causation, and specifically in the context of mediation studies, to measurement error and collider bias^15^.

Mendelian randomisation (MR) is an instrumental variable technique that uses genetic variants as a proxy for an exposure of interest^16^. These variants are randomised at conception and therefore largely inherited independently from other variants affecting confounding factors. These variants are also unchanged throughout the lifetime and so are unlikely to be affected by outcomes of interest, therefore reducing bias from reverse causation. Thus, MR reduces the sources of bias that limit causal inference in conventional observational research and is an effective, complimentary tool in investigating potential causality.

Here, we specifically used two-step MR using summary data for each step. Two-step MR measures the exposure-mediator, exposure-outcome and mediator-outcome effects separately and is particularly useful when binary mediators are being measured individually. We utilised separate samples for the instrument and outcome summary data, thus providing greater sample sizes in some cases.

In this study, we used a complimentary approach utilising both observational and MR techniques to explore the relationship between education, mental health and cardiovascular disease. Elucidation of this causal pathway could have important implications for policymakers interested in both physical and mental health improvement.

## Methods

Data availability: The data used for the observational analyses conducted in this study is available to qualified researchers upon application to the UK biobank (https://www.ukbiobank.ac.uk/researchers/). The data used for the MR analyses conducted in this study is publicly available via the cited references.

### I. Observational analyses

#### Sample

Our observational analyses used data from the UK Biobank, a national health resource which recruited participants aged 39-72 years from 22 centres across the UK between 2006-2010. Details of recruitment are publicly available^17^. We excluded 814 individuals on the basis of consent withdrawal, reported aneuploidy, or mismatch between reported and chromosomal sex. We then applied the Medical Research Council Integrative Epidemiology Unit quality control procedure in order to restrict the sample to those of European ancestry^18^. The final sample size with complete phenotype measures was 333,525 with a mean age of 56.9 years (standard deviation [SD] = 8) and 54% females.

#### Measures

Educational attainment: Participants in the UK Biobank reported the levels of qualifications attained and the highest level of qualification was converted into number of years in education using the International Standard Classification for Education coding of educational attainment (See Supplementary Table S1). Mean educational attainment was 13.9 years (SD = 5.1).

Mental health problems: Individuals were classified as having had a mental health problem if they reported “Yes” to either of the routine survey questions “Have you ever seen a general practitioner (GP) for nerves, anxiety, tension or depression?” or “Have you ever seen a psychiatrist for nerves, anxiety, tension or depression?”.

Depression: Individuals were classified as having major depressive disorder if their hospital episode statistics and/or general practice linked health record showed a diagnosis of a depressive episode or recurrent episodes according to the International Classification of Disease (ICD) codes F32 and F33.

Anxiety: Individuals were classified as having anxiety disorder if their health record showed such a diagnosis according to ICD code F41.

The prevalence of these mediators in our sample was 34% for mental health problems, 2.9% for depression, and 1.4% for anxiety.

*Cardiovascular disease:* Individuals were classified as having cardiovascular disease if their health records showed at least one diagnosis of angina or myocardial infarction prior to recruitment classified by ICD codes I20 and I21 respectively. The prevalence of cardiovascular disease in the sample at the time of recruitment was 6%.

Additional covariates included age, sex, smoking status, hypertension, obesity, and socio-economic position. The methods for their classification and the respective prevalence figures are reported in the supplementary material (see Supplementary Information and Supplementary Table S2).

### II. MR analyses

#### Genetic instruments

Educational attainment: 1271 independent genome-wide significant loci (p<5×10^-8^) were identified in a GWAS meta-analysis of educational attainment by Lee and colleagues (N = 1,131,881)^19^. Educational attainment was measured as years in education. The GWAS meta-analysed 71 cohorts of European ancestry measuring educational attainment. The median effect size of each allele of these lead variants was 1.7 weeks of schooling.

Mental health problems: 14 independent genome-wide significant loci were identified in a recent GWAS of a phenotype described as “broad depression” in UK Biobank data (113,769 cases and 208,811)^20^. Here, we report this phenotype as “Having had a mental health problem” instead. Individuals were classified as having had a mental health problem if they reported “Yes” to either of the routine survey questions “Have you ever seen a general practitioner (GP) for nerves, anxiety, tension or depression?” or “Have you ever seen a psychiatrist for nerves, anxiety, tension or depression?”. Cases with diagnosed bipolar disorder or schizophrenia or who were taking anti-psychotic medications were excluded. A number of participants were also excluded by inter-relatedness filtering.

Depression: 44 genome-wide significant loci were identified in a GWAS meta-analysis of major depressive disorder (135,458 cases and 344,901 controls) by Wray and colleagues^21^. A total of 29 samples containing individuals of European ancestry were included in the analysis. Major depressive disorder was defined by meeting DSM-IV or IC9-9/10 criteria either by clinician review of medical records or by expert interview.

Anxiety: A recent GWAS of anxiety disorder (7016 cases and 14,745 controls) identified one independent genome-wide significant locus^22^. Anxiety cases were recruited from seven different cohorts of European ancestry participating in the Anxiety NeuroGenetics Study with cases confirmed according to DSM-IV criteria. In order to improve instrument power, we relaxed the significance threshold to p<5×10^-5^ and identified 108 independent significant loci following LD clumping (r2<0.001, distance>10000 kb).

Cardiovascular disease: We used summary data from a recent GWAS meta-analysis from the CardioGRAMplusC4D consortium^23^. This included 48 cohorts with a total of 60,801 cases and 123,504 controls. The cohort studies typically defined a case as having a diagnosis of myocardial infarction or angina with several also requiring confirmation from angiographic evidence.

### III. Statistical analysis

All analyses were performed using R (Version 3.6.2)^24^. A directed acyclic graph for the proposed causal relationships investigated is shown in Figure 1.

**Figure 1.**
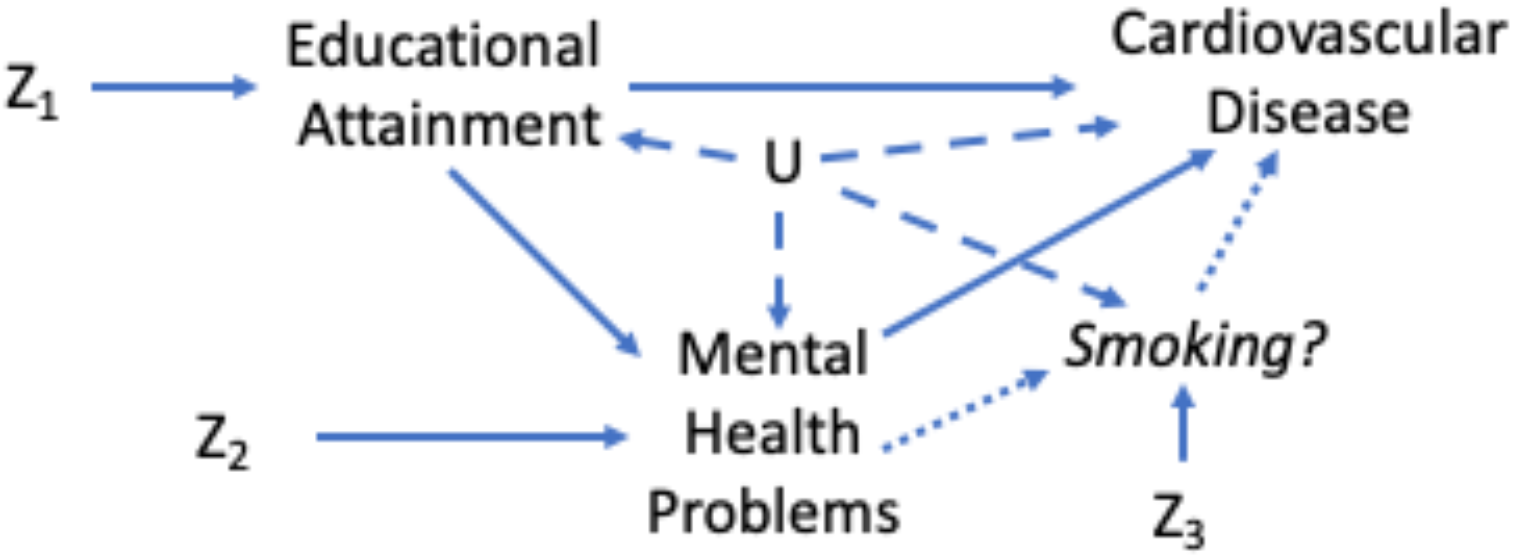
*A diagram illustrating the proposed causal relationships between education, mental health and cardiovascular disease. Z_1_ and Z_2_ represent genetic proxies and U represents confounding factors*.

Observational analysis: First, we used logistic regression to investigate the association of educational attainment with cardiovascular disease and with each of our mediators having adjusted for the effects of age and sex. Second, we analysed the effect of each of our mediators on cardiovascular disease and, in order to effectively isolate the mediator effects, adjusted for age, sex, educational attainment, smoking status, socio-economic position, hypertension, and obesity. When specifically analysing the effects of depression on cardiovascular disease, we additionally adjusted for the effects of anxiety and vice versa.

The proportion mediated statistic was calculated using the product of coefficients method as detailed in previous work^25^. The effect sizes were calculated using absolute risk differences as estimated from the odds ratios from our regression analysis and the known prevalence figures in UK Biobank. Standard errors were calculated using the delta method.

MR univariable analysis: The univariable analysis consisted of using two-sample MR methods applied in the TwoSample MR package 0.4.18^26^. We first ran our analysis using educational attainment as an exposure and each of our proposed mediators and cardiovascular disease as outcomes. We then repeated the analysis using each mediator as an exposure and cardiovascular disease as an outcome.

For these analyses, we used an inverse-variance weighted (IVW) approach in the main analysis ^27^, with additional statistical sensitivity analyses: weighted mode^28^, weighted median^29^, and MR Egger^30^. Each sensitivity analysis makes different assumptions about pleiotropy.

When using the anxiety disorder instrument, we performed MR Robust Associated Profile Score (MR RAPS) instead of IVW to improve estimation whilst using a relaxed p-value threshold for the instrument^31^. MR RAPS offers a greater robustness to pleiotropy when using many weak instruments, as we do here.

Two-step MR analysis: We next performed two-step MR using the TwoSample MR package 0.4.18. We used the results of our univariable MR analysis for the exposure-mediator step. For the mediator-outcome step we then used the residual multivariable MR method to calculate the effect of each mediator on cardiovascular disease having adjusted for the effects of educational attainment^32^. The estimates for each step were multiplied together to produce an indirect effect estimate and the proportion mediated statistic was then calculated as for the observational analysis (detailed above).

### IV. Sensitivity analyses

The methods for our sensitivity analyses are presented in the supplementary material (see Supplementary Information). These analyses included MR Egger, Cochran’s Q Tests of Heterogeneity, mean of F (mF) statistics, MR Steiger, and bidirectional observational models. They also included an observational analysis of mental health and cardiovascular disease conducted without adjustment for other co-variates except for age and sex.

### V. Exploratory analysis

In order to illustrate an approach for investigating multiple mediators between mental health and cardiovascular disease, we performed an exploratory analysis of a likely candidate based on existing evidence. This analysis investigates the role of tobacco smoking as a mediator between depression and cardiovascular disease. The methods are in Supplementary Material.

Ethical Considerations: UK Biobank has received ethics approval from the UK National Health Service’s National Research Ethics Service (ref 11/NW/0382).

## Results

### Association of education with cardiovascular disease

We found strong evidence to suggest that educational attainment has an inverse association with cardiovascular disease (see Figure 2). In our observational analysis, for every standard deviation increase in educational attainment there was a 24% (95% CI 21-27%) reduction in cardiovascular disease. Our MR analysis suggested a causal, protective effect for education against cardiovascular disease.

### Association of education with mental health

We also found that education has an inverse association with anxiety and depression and that this is consistent across both adjusted observational and MR analyses (see Figure 2). For each standard deviation increase in educational attainment there were 25% (95% CI 24-26%) and 21% (95% CI 19-23%) reductions in depression and anxiety respectively. Our MR analysis suggested that education has a causal, protective effect against anxiety and depression.

Educational attainment generally showed only weak evidence of an association with mental health problems in our observational analysis (OR 0.99, 95% CI 0.98-1.01, p=0.06) but, in our MR analysis, there was evidence of a causal, protective effect (IVW OR 0.95, 95% CI 0.94-0.96, p<0.0001). This association was consistent across the MR sensitivity analyses.

**Figure 2.**
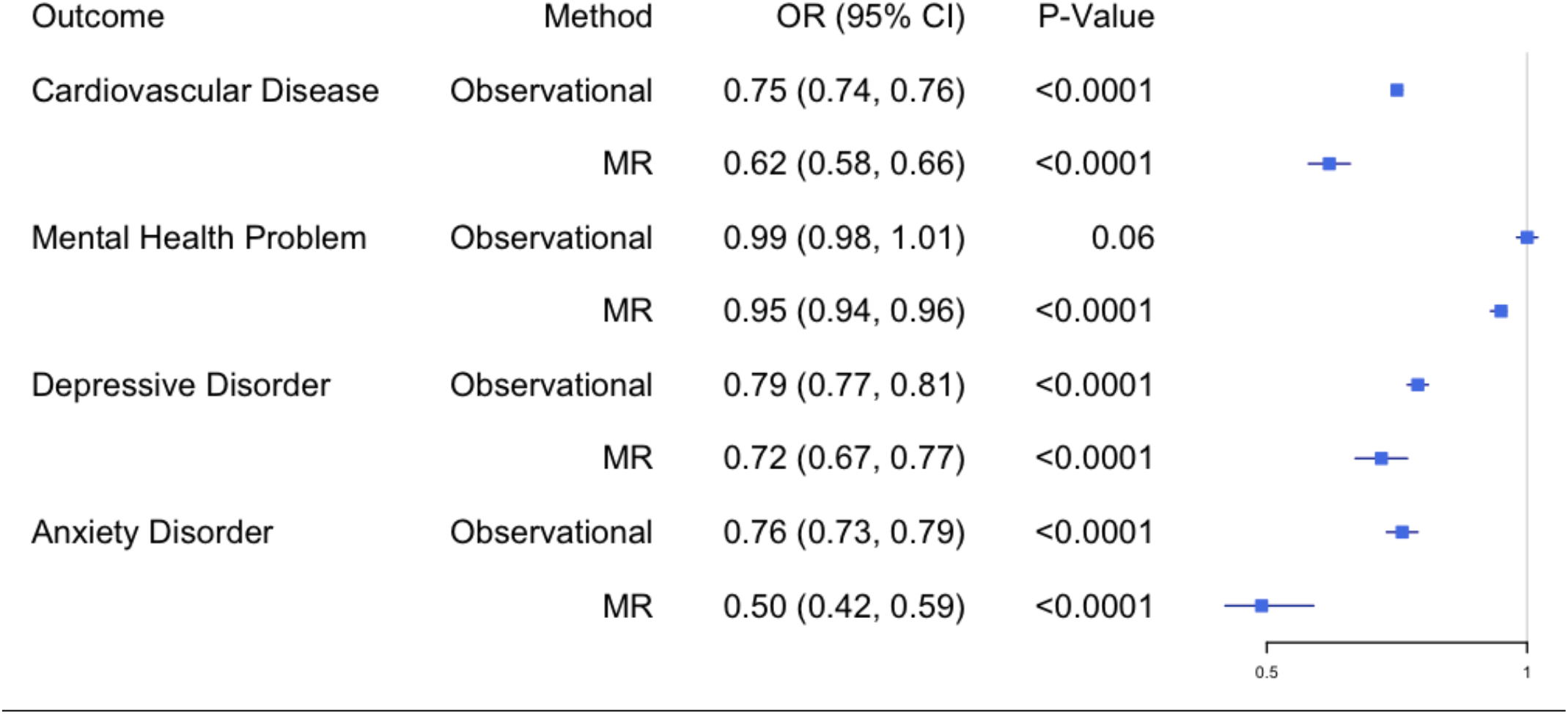
*The association of educational attainment with mental health and cardiovascular disease presented as odds ratios per standard deviation increase in education*

### Association of mental health with cardiovascular disease

In our observational analysis, mental health problems in general were associated with increased cardiovascular disease risk (OR 1.29, 95% CI 1.24-1.33, p<0.001), but our MR analysis did not provide clear support for a causal interpretation (OR 1.48, 95% CI 0.92-2.39, p=0.11) (see Figure 3).

However, we found strong observational evidence that depression was associated with increased cardiovascular risk (OR 1.70, 95% CI 1.58-1.82, p<0.001), and our MR analyses supported a causal interpretation (OR 1.09, 95% CI 1.03-1.15, p=0.0021). Anxiety disorder was strongly associated with cardiovascular disease in our observational analysis (OR 1.63, 95% CI 1.49-1.79, p<0.001) but MR analysis did not support a causal interpretation (OR 1.00, 95% CI 0.99-1.01, p=0.82).

**Figure 3.**
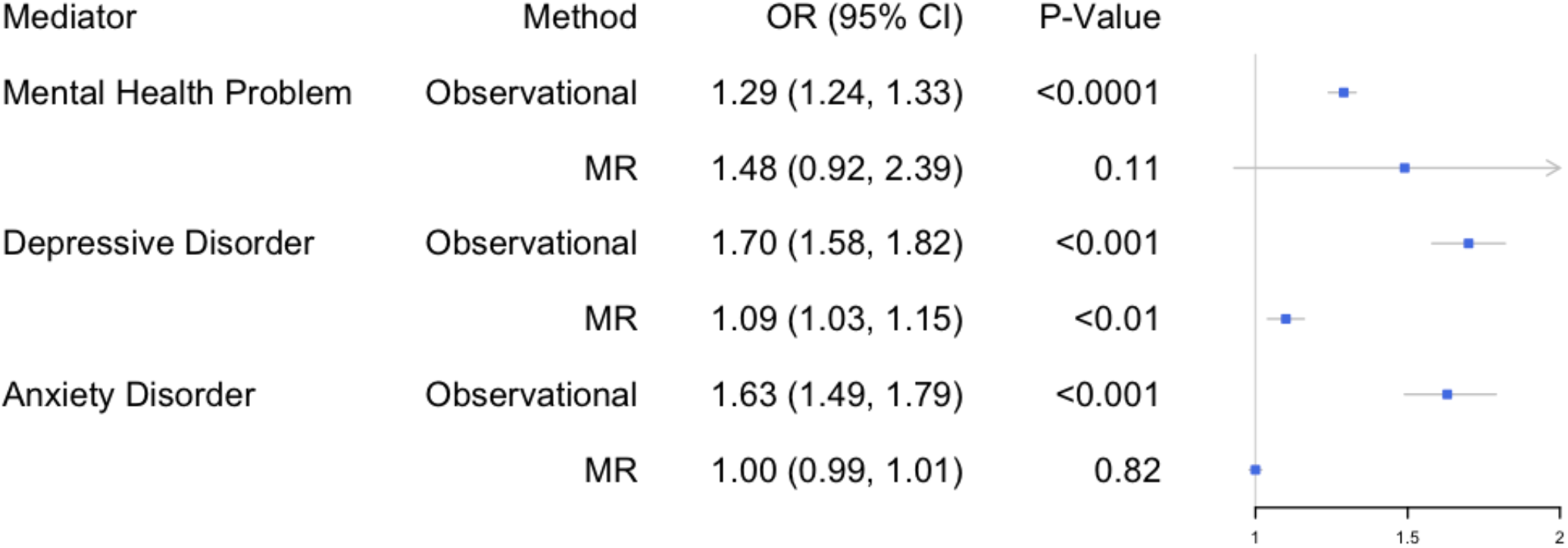
*The association of mental health with cardiovascular disease following adjustment for the effects of educational attainment*

### Proportion mediated by mental health of the association between education and cardiovascular disease

Our mediation results show that mental health problems in general (2-step MR: 4.2%, 95% CI 3.0%-5.3%) and specifically depression (2-step MR: 6.5%, 95% CI 5.6%-7.5%) account for a small proportion of the negative association between education and cardiovascular disease (see Figure 4). The proportion mediated by anxiety disorder was very small in the observational analysis, and we found little evidence of mediation in our MR analysis.

**Figure 4.**
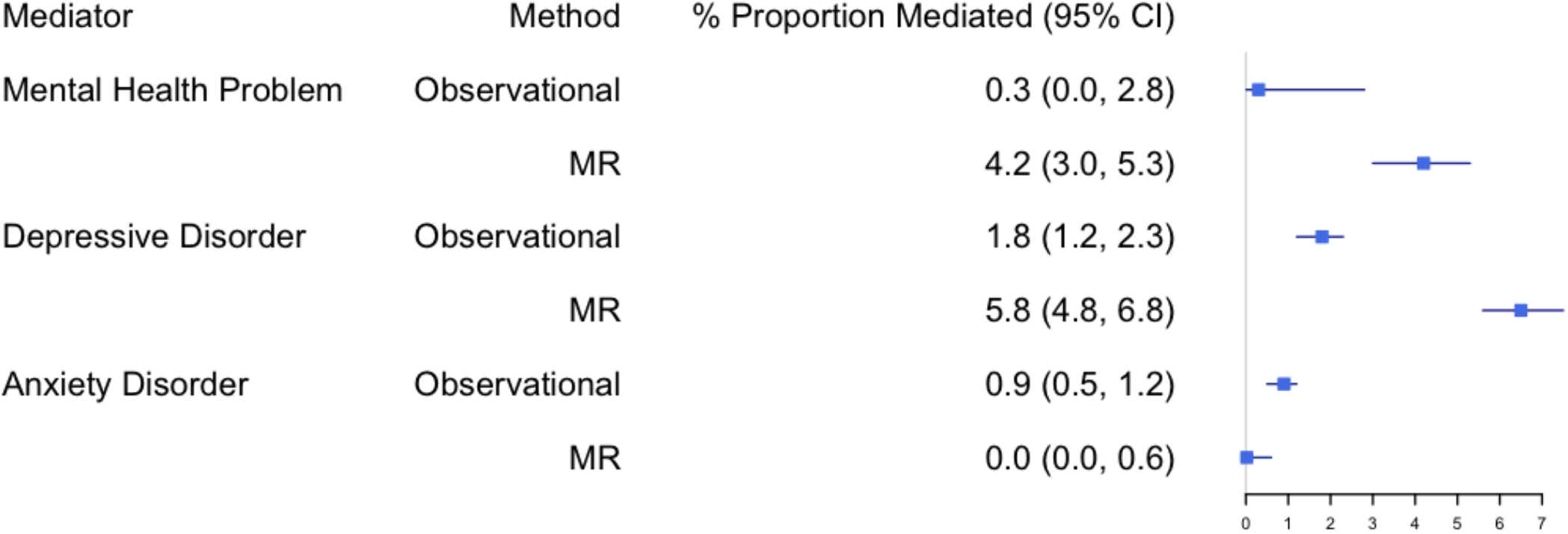
*The estimated proportions of the inverse association between education and cardiovascular disease accounted for by mental health presentations, depression and anxiety*

### Sensitivity analyses

The results of our sensitivity analyses are presented in Supplementary Tables S3-13 and included: testing for pleiotropy and reverse causation, testing for bidirectionality in our observational data, and testing personality traits as additional potential mediators. The results provided little evidence of bias from directional pleiotropy but did raise the possibility of reverse causation between cardiovascular disease and the proposed mediators in our observational analyses.

### Exploratory analysis of tobacco smoking as a mediator between depression and cardiovascular disease

In a further analysis, we sought to investigate a potential role for multiple mediators in this pathway, specifically smoking as a mediator between adjusted depression and cardiovascular disease. We found that depression is associated with increased smoking activity and that lifetime smoking was associated with increased cardiovascular risk (see Supplementary Tables S15-16). In Figure 5, we show that smoking accounts for a large proportion of the increased cardiovascular risk associated with depression, 29% (95% CI 28.9-29.7%) according to our MR analysis.

**Figure 5.**
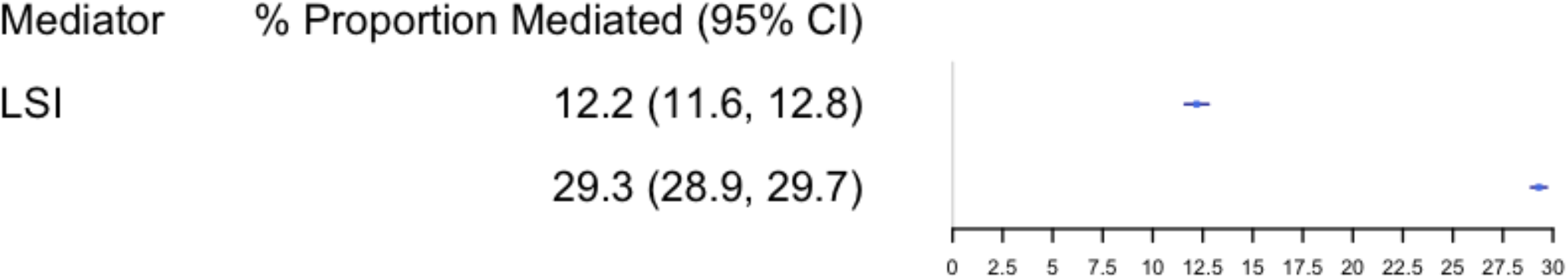
*The estimated proportion of the association between depression and cardiovascular disease accounted for by lifetime smoking*

## Discussion

In this study, we used both observational and MR techniques to investigate mental health as a mediator between education and cardiovascular disease. Our results demonstrate that depression accounts for a proportion of the protective effect education has on cardiovascular disease.

We demonstrated, in observational analyses, that educational attainment has a clear, inverse association with anxiety disorder, major depressive disorder, and cardiovascular disease. Our MR analyses suggested these relationships are causal. In addition, our MR analyses suggested a causal, protective effect of educational attainment on mental health problems more broadly although observational analysis showed no strong association.

Mental health problems in general and, specifically, anxiety and depression were all associated with increased cardiovascular risk in our observational analyses. Our MR analyses only suggested a causal relationship between depression and cardiovascular disease.

Following these initial results, we further explored the proposed causal relationships between education, mental health and cardiovascular disease, which likely consists of additional mediators (see Figure 1). Our observational analyses showed that depression was positively associated with lifetime smoking which was, in turn, associated with increased cardiovascular risk even after adjustment for depression. Our MR analyses suggested these associations are causal. We found that lifetime smoking accounts for as much as 30% of the association between depression and cardiovascular disease.

### Our results in context

Our analysis suggests that depression serves as a mediator between education and cardiovascular disease, and thus accounts for an unexplained portion of the protective effect of education. Specifically, it suggests that educational disparities may produce additional cardiovascular disease burden partly by increasing rates of depression. Depression and coronary artery disease have previously been found to be associated in MR work^33,34^; however, this mediation analysis constitutes a novel finding that can help identify possible new targets for interventions.

Our results support a protective role for education against mental health problems and, specifically, major depressive disorder and anxiety disorder. The foremost explanation for this is educational attainment being a proxy for overall socio-economic position which has a robust negative correlation with depression and anxiety^35^. For example, educational attainment is associated with both higher income and better occupation later in life ^36,37^. Such factors protect against experiences such as debt, financial struggles, and poor working conditions which feature amongst several relevant risk factors for mental health disorders^38^. Related interventions would accordingly include policies to promote educational attainment and reduce socio-economic inequalities.

At an individual level, further years spent in education or training are also likely to play a protective role against mental health problems by directly modifying a range of psychosocial factors and providing resources that can be drawn upon as stressors are encountered^39^. Such resources might include mental health advice, support networks, social participation, close friends, and good work-life balance^40,41^. Additional years in education may also help to offset exposure to stressors until one has greater maturity and resources to cope with them. By considering these observations, successful individual-level interventions in the education system might be developed.

In an exploratory analysis, we found that smoking accounts for a large proportion of the association between depression and cardiovascular disease. The link between smoking and cardiovascular disease is well-known and depression and smoking are also associated^42,43^. This link between smoking and depression has been commonly attributed to a ‘self-medication’ hypothesis where smoking is used to cope with depressive symptoms. Other factors that are proposed to partially explain the remainder of the association between depression and cardiovascular disease include chronic inflammation and changes in lipid metabolism^44^.

### Implications

Our study has several important public health implications. First, it strengthens the case for investment in education and interventions related to it by demonstrating that educational attainment can reduce mental health burden, which is itself a significant source of morbidity and mortality^45^. Second, by illustrating the links between education, mental health, and cardiovascular disease, we have reinforced the need to recognise the potential interplay between social determinants, physical and mental health problems. Indeed, in this study, we show how a physical disease burden could be partly alleviated by targeting a social determinant of mental health. Finally, we have also illustrated how to further explore potential mediators between depression and cardiovascular disease in order to elucidate additional intervention targets. Here, we have highlighted tobacco smoking as such a target.

### Strengths and limitations

The strengths of our study include the use of MR analysis to complement a conventional observational approach which allowed us to better overcome bias arising from confounding, measurement error and reverse causation. The use of a ‘triangulation approach’ also allowed for greater confidence in causal inference when there was consistency between the different approaches^46^. We were also able to limit bias arising from self-reporting and misclassification by using strict case definition for both our observational and MR data sources. Such bias can be particularly problematic when using binary exposures in MR^47^. An additional strength of our study was the use of two-step MR for a mediation analysis. This allowed us to specifically quantify the proportion of our exposure’s effect attributable to our proposed mediators. Finally, we extended our analysis in order to illustrate how our methodology could explore multiple mediators in the proposed pathway.

In our study, the use of MR demonstrated a discrepancy when observing the effects of mental health on cardiovascular disease. When adjusting for confounding in our mediation analysis, we likely over-adjusted in order to confidently isolate the effects of the mediator. To measure this, we conducted a sensitivity analysis without adjustment although this suggested that the effect sizes we have found may actually be underestimates (see Supplementary Table 17). Instead, the discrepancy may have primarily been due to reverse causation, with cardiovascular disease leading to mental health problems and exaggerating observed associations. In a further sensitivity analysis, where cardiovascular disease was the exposure and depression and then anxiety the outcomes, we did indeed find such bidirectionality.

A common limitation of MR studies is weak instrument strength^48^. Our instruments for both educational attainment and depression were adequate in this regard, with large mF statistics and sample sizes. However, our instrument for anxiety had a relatively smaller mF statistic and required a relaxed genome-wide significance threshold; the null result should therefore be interpreted with caution. However, we utilised the MR RAPS method to compensate for the relaxed threshold and we also conducted a sensitivity analysis with the worry sub-scale of the Eysenck Personality Questionnaire (Revised Short form). This analysis had adequate instrument strength and a larger sample size but also produced a null result (see Supplementary Table S8).

Despite adequate instrument strength, our ‘mental health problems’ instrument produced wide confidence intervals, suggesting these analyses may have been underpowered. Unlike our other instruments, this one was constructed from a GWAS performed on a single sample, as the unusual phenotype measured is only available in the UK Biobank. In future, we hope other cohorts will have this or a similar phenotype measured, leading to larger sample sizes for analysis and, hopefully, the discovery of further significant variants.

When conducting two-sample MR, overlap between the samples can be problematic. The samples used for our exposure and mediators did feature partial overlap due to the use of UK Biobank data in both sets. This introduces a greater risk of a type I error in our two-sample univariable analyses of education and mental health due to possible overfitting. However, we also saw an inverse association with education in our sensitivity analyses that utilised personality traits related to mental health (see Supplementary Table S8). These featured substantively less overlap between samples. In addition, this limitation did not apply to our two-step analysis which pools the data for the exposure and mediators before analysis is conducted.

Another common limitation of MR studies is bias from directional pleiotropy^49^. We conducted numerous sensitivity analyses, including multiple MR methods and measurement of MR Egger intercepts (see Supplementary Tables S3 and S7), which suggest our results were not biased by directional pleiotropy.

Finally, even with our approach, education may still be acting as a proxy for socioeconomic position. A proposed strategy for investigating this would be to conduct an additional analysis using a measure such as household income which would allow for comparison as well as adjustment as part of a multivariable MR approach. A recently published GWAS meta-analysis exploring household income^50^ offers the possibility of this in future studies. Natural experiment techniques could also be used having previously suggested that the raising of school age reduced cardiovascular disease^51^.

### Conclusions

In conclusion, we have demonstrated that educational attainment is inversely associated with mental health problems, anxiety, depression, and cardiovascular disease. A reduction in depression accounts for part of the inverse association between education and cardiovascular disease. Finally, we have shown that smoking accounts for a large proportion of the association between depression and cardiovascular disease. Our findings offer significant clinical and public health implications by further demonstrating interplay between social determinants, mental and physical disease and associated targets for intervention.

## Data Availability

The data used for the observational analyses conducted in this study is available to qualified researchers upon application to the UK biobank (https://www.ukbiobank.ac.uk/researchers/). The data used for the MR analyses conducted in this study is publicly available via the cited references.

## Acknowledgements

We are grateful to the participants of the UK Biobank and the individuals who contributed to each of the previous GWAS analyses conducted as well as all the research staff who worked on the data collection. This research has been conducted using the UK Biobank Resource under Application Number 9142.

## Supplementary Materials

Supplementary Methods

Supplementary Tables S1-S15

Supplementary Figures S16-S17

References 52-571

## References

1 Global atlas on cardiovascular disease prevention and control. WHO. https://www.who.int/cardio-vascular_diseases/publications/atlas_cvd/en/. Published January 2020. Accessed January 25, 2020.

2 Inequalities in health. GOV.UK. https://www.gov.uk/government/publications/health-profile-for-england-2018/chapter-5-inequalities-in-health. Published 2018. Accessed January 25, 2020.

3 Rosengren A, Subramanian S, Islam S, Chow C, Avezum A, Kazmi K, Sliwa K, Zubaid M, Rangarajan S, Yusuf S. Education and risk for acute myocardial infarction in 52 high, middle and low-income countries: INTERHEART case-control study. Heart. 2009;95(24):2014–2022.

4 Tillmann T, Vaucher J, Okbay A, Pikhart H, Peasey A, Kubinova R, Pajak A, Tamosiunas A, Malyutina S, Hartwig F, Fischer K, Veronesi G, Palmer T, Bowden J, Davey Smith G et al. Education and coronary heart disease: mendelian randomisation study. BMJ. 2017:j3542.

5 Lynch J, Kaplan G, Cohen R, Tuomilehto J, Salonen J. Do Cardiovascular Risk Factors Explain the Relation between Socioeconomic Status, Risk of All-Cause Mortality, Cardiovascular Mortality, and Acute Myocardial Infarction? American Journal of Epidemiology. 1996;144(10):934–942.

6 Carter A, Gill D, Davies N, Taylor A, Tillmann T, Vaucher J, Wootton R, Munafò M, Hemani G, Malik R, Seshadri S, Woo D, Burgess S, Davey Smith G, Holmes M et al. Understanding the consequences of education inequality on cardiovascular disease: mendelian randomisation study. BMJ. 2019:l1855.

7 Hemingway H, Marmot M. Evidence based cardiology: Psychosocial factors in the aetiology and prognosis of coronary heart disease: systematic review of prospective cohort studies. BMJ. 1999;318(7196):1460–1467.

8 Melkevik O, Hauge L, Bendtsen P, Reneflot A, Mykletun A, Aara L. Associations between delayed completion of high school and educational attainment and symptom levels of anxiety and depression in adulthood. BMC Psychiatry. 2016;16(1).

9 Gallo L, Matthews K. Understanding the association between socioeconomic status and physical health: Do negative emotions play a role? Psychological Bulletin. 2003;129(1):10–51.

10 Lazzarino A, Hamer M, Gaze D, Collinson P, Rumley A, Lowe G, Steptoe A. The interaction between systemic inflammation and psychosocial stress in the association with cardiac troponin elevation: A new approach to risk assessment and disease prevention. Preventive Medicine. 2016;93:46–52.

11 Hering D, Lachowska K, Schlaich M. Role of the Sympathetic Nervous System in Stress-Mediated Cardiovascular Disease. Current Hypertension Reports. 2015;17(10).

12 Iob E, Steptoe A. Cardiovascular Disease and Hair Cortisol: a Novel Biomarker of Chronic Stress. Current Cardiology Reports. 2019;21 (10).

13 Matthews K, Gallo L, Taylor S. Are psychosocial factors mediators of socioeconomic status and health connections? Annals of the New York Academy of Sciences. 2010;1186(1):146–173.

14 Boylan J, Cundiff J, Matthews K. Socioeconomic Status and Cardiovascular Responses to Standardized Stressors. Psychosomatic Medicine. 2018;80(3):278–293.

15 Alice R Carter, Eleanor Sanderson, Gemma Hammerton, Rebecca C Richmond, George Davey Smith, Jon Heron, Amy E Taylor, Neil M Davies, Laura D Howe. (Preprint) Mendelian randomisation for mediation analysis: current methods and challenges for implementation. bioRxiv. 835819. doi: https://doi.org/10.1101/835819

16 Baiocchi M, Cheng J, Small D. Instrumental variable methods for causal inference. Statistics in Medicine. 2014;33(13):2297–2340.

17 Sudlow C, Gallacher J, Allen N, Beral V, Burton P, Danesh J, Downey P, Elliott P, Green J, Landray M, Liu B, Matthews P, Ong G, Pell J, Silman A et al. UK Biobank: An Open Access Resource for Identifying the Causes of a Wide Range of Complex Diseases of Middle and Old Age. PLOS Medicine. 2015;12(3):e1001779.

18 Mitchell R, Hemani G, Dudding T, Corbin L, Harrison S, Paternoster L. IEU Quality Control. Data.bris.ac.uk. https://www.r-pro. Published 2019. Accessed February 24, 2020.

19 Lee J, Wedow R, Okbay A, Kong E, Maghzian O, Zacher M, Nguyen-Viet T, Bowers P, Sidorenko J, Karlsson Linnér R, Fontana M, Kundu T, Lee C, Li H, Li R et al. Gene discovery and polygenic prediction from a genome-wide association study of educational attainment in 1.1 million individuals. Nature Genetics. 2018;50(8):1112–1121.

20 Howard D, Adams M, Shirali M, Clarke T, Marioni R, Davies G, Coleman J, Alloza C, Shen X, Barbu M, Wigmore E, Gibson J, Hagenaars S, Lewis C, Ward J et al. Genome-wide association study of depression phenotypes in UK Biobank identifies variants in excitatory synaptic pathways. Nature Communications. 2018;9(1).

21 Wray N, Ripke S, Mattheisen M, Trzaskowski M, Byrne E, Abdellaoui A, Adams M, Agerbo E, Air T, Andlauer T, Bacanu S, Bækvad-Hansen M, Beekman A, Bigdeli T, Binder E et al. Genome-wide association analyses identify 44 risk variants and refine the genetic architecture of major depression. Nature Genetics. 2018;50(5):668–681.

22 Otowa T, Hek K, Lee M, Byrne E, Mirza S, Nivard M, Bigdeli T, Aggen S, Adkins D, Wolen A, Fan-Fan-ous A, Keller M, Castelao E, Kutalik Z, der Auwera S et al. Meta-analysis of genome-wide association studies of anxiety disorders. Molecular Psychiatry. 2016;21(10):1391–1399.

23 The CARDIoGRAMplusC4D Consortium. A comprehensive 1000 Genomes-based genome-wide association meta-analysis of coronary artery disease. Nature Genetics. 2015;47(10):1121–1130.

24 R Core Team. R: The R Project for Statistical Computing. R-project.org. https://www.r-pro-ject.org/. Published 2014. Accessed January 26, 2020.

25 VanderWeele TJ. Mediation Analysis: A Practitioner’s Guide. Annual Review Public Health. 2016; 37: 17–32

26 MR Base for two sample MR. MRCIEU. https://mrcieu.github.io/TwoSampleMR/. Published 2019. Accessed January 27, 2020.

27 Lawlor D, Harbord R, Sterne J, Timpson N, Davey Smith G. Mendelian randomization: Using genes as instruments for making causal inferences in epidemiology. Statistics in Medicine. 2008;27(8):1133–1163.

28 Hartwig F, Davey Smith G, Bowden J. Robust inference in summary data Mendelian randomization via the zero modal pleiotropy assumption. International Journal of Epidemiology. 2017; 46(6):1985–98.

29 Bowden J, Davey Smith G, Haycock P, Burgess S. Consistent Estimation in Mendelian Randomization with Some Invalid Instruments Using a Weighted Median Estimator. Genetic Epidemiology. 2016; 40(4):304–14.

30 Burgess S, Thompson S. Interpreting findings from Mendelian randomization using the MR-Egger method. European Journal of Epidemiology. 2017; 32(5):377–89.

31 Qingyuan Z, Jingshu W, Jack B, Dylan SS. Statistical inference in two-sample summary-data Mendelian randomization using robust adjusted profile score. (Preprint) Arvix. 2018; https://arxiv.org/abs/1801.09652

32 Burgess S, Daniel RM, Butterworth AS, Thompson SG; EPIC-InterAct Consortium. Network Mendelian randomization: using genetic variants as instrumental variables to investigate mediation in causal pathways. International Journal of Epidemiology. 2015; 44:484–95.

33 Wu Q, Kling JM. Depression and the risk of myocardial infarction and coronary death: a metaanalysis of prospective cohort studies. Medicine. 2016; 95:e2815.

34 Mulugeta A, Zhou A, King C, Hyppönen E. Association between major depressive disorder and multiple disease outcomes: a phenome-wide Mendelian randomisation study in the UK Biobank. Molecular Psychiatry. 2019;25(7):1469–1476.

35 Fryers T, Melzer D, Jenkins R. Social inequalities and the common mental disorders. Social Psychiatry and Psychiatric Epidemiology. 2003;38(5):229–237.

36 Education and earnings. OECD. https://stats.oecd.org/Index.aspx?DataSetCode=EAG_EARNINGS. Published 2020. Accessed on 28 January 2020

37 Andersen R, Van De Werfhorst H. Education and occupational status in 14 countries: the role of educational institutions and labour market coordination. British Journal of Sociology. 2010; 61 (2):336–55.

38 Weich S, Lewis G. Poverty, unemployment and the common mental disorders: a population-based cohort study. BMJ. 1998; 317:115–19.

39 Matthews KA, Gallo LC. Psychological perspectives on pathways linking socioeconomic status and physical health. Annual Review of Psychology. 2011; 62:501–30

40 Aldabe B, Anderson R, Lyly-Yrjanainen M, Parent-Thirion A, Vermeylen G, Kelleher C, Niedhammer I. Contribution of material, occupational, and psychosocial factors in the explanation of social inequalities in health in 28 countries in Europe. Journal of Epidemiology & Community Health. 2010; 65(12):1123–1131.

41 Moor I, Rathmann K, Stronks K, Levin K, Spallek J, Richter M. Psychosocial and behavioural factors in the explanation of socioeconomic inequalities in adolescent health: a multilevel analysis in 28 European and North American countries. Journal of Epidemiology and Community Health. 2014; 68(10):912–921.

42 Doll R, Peto R, Boreham J, Sutherland I. Mortality in relation to smoking: 50 years’ observations on male British doctors. BMJ. 2004; 328:1519

43 Taylor A, Fluharty M, Munafò M. Investigating the possible causal association between smoking and depression and anxiety using Mendelian randomisation meta-analysis: The CARTA consortium. Journal of Epidemiology and Community Health. 2014; 68(Suppl 1):A45.1-A45.

44 Khandaker G, Zuber V, Rees J, Carvalho L, Mason A, Foley C, Gkatzionis A, Jones P, Burgess S. Shared mechanisms between coronary heart disease and depression: findings from a large UK general population-based cohort. Molecular Psychiatry. 2019;25(7):1477–1486.

45 Mathers CD, Loncar D. Projections of Global Mortality and Burden of Disease from 2002 to 2030. PLoS Medicine. 2006; 3(11):e442.

46 Lawlor D, Tilling K, Davey-Smith G. Triangulation in aetiological epidemiology. International Journal of Epidemiology. 2016; 45(6): 1866–1886.

47 Burgess S, Labrecque JA. Mendelian randomization with a binary exposure variable: interpretation and presentation of causal estimates. European Journal of Epidemiology. 2018; 33: 947–952.

48 Davies Neil M, Holmes Michael V, Davey-Smith G. Reading Mendelian randomisation studies: a guide, glossary, and checklist for clinicians BMJ. 2018; 362:k60.

49 Hemani G, Bowden J, Davey-Smith G. Evaluating the potential role of pleiotropy in Mendelian randomization studies. Human Molecular Genetics. 2018; 27(R2):R195–R208.

50 Hill W, Davies N, Ritchie S, Skene N, Bryois J, Bell S, Di Angelantonio E, Roberts D, Xueyi S, Davies G, Liewald D, Porteous D, Hayward C, Butterworth A, McIntosh A et al. Genome-wide analysis identifies molecular systems and 149 genetic loci associated with income. Nature Communications. 2019; 10(1):5741.

51 Hamad R, Nguyen TT, Bhattacharya J, Glymour MM, Rehkopf DH. Educational attainment and cardiovascular disease in the United States: A quasi-experimental instrumental variables analysis. PLoS Medicine. 2019; 16(6):e1002834.

52 Bowden J, Del Greco M. F, Minelli C, Davey-Smith G, Sheehan N, Thompson J. Assessing the suitability of summary data for two-sample Mendelian randomization analyses using MR-Egger regression: the role of the I2 statistic. International Journal of Epidemiology. 2016: dyw220.

53 Brion M, Shakhbazov K, Visscher P. Calculating statistical power in Mendelian randomization studies. International Journal of Epidemiology. 2012; 42(5):1497–1501.

54 Greco M FD, Minelli C, Sheehan NA, Thompson JR. Detecting pleiotropy in Mendelian randomisation studies with summary data and a continuous outcome. Statistics in Medicine. 2015; 34:2926–40.

55 Hemani G, Tilling K, Davey Smith G. Orienting the causal relationship between imprecisely measured traits using GWAS summary data. PloS Genetics. 2017;13(11):e1007081.

56 Nagel M, Jansen P, Stringer S, Watanabe K, de Leeuw C, Bryois J, Savage J, Hammerschlag A, Skene N, Muñoz-Manchado A, White T, Tiemeier H, Linnarsson S, Hjerling-Leffler J, Polderman T et al. Meta-analysis of genome-wide association studies for neuroticism in 449,484 individuals identifies novel genetic loci and pathways. Nature Genetics. 2018; 50(7):920–927.

57 Liu M, Jiang Y, Wedow R, Li Y, Brazel D, Chen F, Datta G, Davila-Velderrain J, McGuire D, Tian C, Zhan X, Choquet H, Docherty A, Faul J, Foerster J et al. Association studies of up to 1.2 million individuals yield new insights into the genetic etiology of tobacco and alcohol use. Nature Genetics. 2019;51(2):237–244.

